# WHAT FUELS SUBOPTIMAL CARE OF PERIPHERAL INTRAVENOUS CATHETER-RELATED INFECTIONS IN HOSPITALS? – A QUALITATIVE STUDY OF DECISION-MAKING AMONG SPANISH NURSES

**DOI:** 10.1101/2021.08.29.21262400

**Authors:** Ian Blanco-Mavillard, Enrique Castro-Sánchez, Gaizka Parra-García, Miguel Ángel Rodríguez-Calero, Miquel Bennasar-Veny, Ismael Fernández-Fernández, Harri Lorente-Neches, Joan de Pedro-Gómez

**Affiliations:** Hospital Manacor, Manacor, Spain; City, University of London, London, United Kingdom; Hospital Universitario Son Llatzer, Palma, Spain; Servei de Salut de les Illes Balears, Palma, Spain; Universitat de les Illes Balears, Palma, Spain; Health Research Institute of the Balearic Islands (IdISBa), Palma, Spain; Care, Chronicity and Evidence in Health Research Group (CurES), Health Research Institute of the Balearic Islands (IdISBa), Palma, Spain

**Keywords:** Clinical decision making, Peripheral Venous Catheterization, Catheter-related infections

## Abstract

**Background:** Peripheral intravenous catheters (PIVC) are commonly used in hospital worldwide. However, PIVC are not exempt from complications. Catheter-related bloodstream infections (CRBSI) increase morbidity and mortality rates, and costs for the healthcare organization. PIVC care is shaped by the complex mix of professional and organizational culture, such as knowledge gaps, low perception of impact of PIVCs on patient safety, or lack of hospital guidelines.

**Aim:** To explore determinants of decision-making about the prevention of PIVC-BSI among nurses in Spanish hospitals.

**Methods:** We conducted a descriptive qualitative study with semi-structured interviews in three public hospitals, the Balearic Islands Health Care Service in Spain. We considered hospital ward nurses working routinely with inpatients at any of the three hospitals for enrolment in the study. We approached relevant informants to identify suitable participants who recruited other participants through a ‘snowball’ technique. Fourteen inpatient nurses from the hospital took part in this study between September and November 2018. We employed several triangulation strategies to underpin the methodological rigour of our analysis and conducted the member checking, showing the information and codes applied in the recording of the interviews to identify the coherence and any discrepancies of the discourse by participants. We used the COREQ checklist for this study.

**Findings:** We identified four major themes in the analysis related to determinants of care: The fog of decision-making in PIVC; The taskification of PIVC care; PIVC care is accepted to be suboptimal, yet irrelevant; and PIVC care gaps may reflect behavioural shortcomings, yet solutions proposed to involve education and training.

**Conclusion:** The clinical management of PIVCs appear ambiguous, unclear, and fragmented, with no clear professional responsibility and no nurse leadership, causing a gap in preventing infections. Furthermore, the perception of low risk on PIVC care impact can cause a relevant lack of adherence to the best evidence and patient safety. Implementing facilitation strategies could improve the fidelity of the best available evidence regarding PIVC care and raise awareness among nurses of impact that excellence of care.

## BACKGROUND

Nurses in hospitals worldwide frequently use peripheral intravenous catheters (PIVCs) [1–3]. In common with virtually all other clinical interventions, the use of PIVCs can result in complications and adverse outcomes for patients, with catheter-related bloodstream infections (CRBSI) one of the worse of these adverse events [4]. As seen in PIVC failure [5,6], healthcare organisations can incur unnecessary expenses and waste resources, fostering dissatisfaction among healthcare professionals and impoverishing the experience of care for patients [7,8], who are subject to increased hospital length of stays, morbidity and mortality [9].

The need for nurses to optimise the management of peripheral intravenous catheter-related bloodstream infections (PIVC-BSI) has already been extensively documented [10–13]. However, these studies did not provide insights into any institutional mechanisms likely in place to assure patient safety and quality in PIVC care [14]. The determinants of optimal catheter use can exert an influence at different levels: individual (i.e., often reported gaps in knowledge and skills among nurses [15]), social (i.e., collective perceptions shaping the relative importance apportioned to PIVCs and, by extension, their adverse events [16]) and, finally, organisational (i.e., unclear guidelines [17] or as our group has reported, lack of patient involvement in their self-care [18,19]).

Healthcare workers must appraise and negotiate the effect of such determinants of practice, ideally using existing evidence. Such evidence, operationalized in clinical practice guidelines (CPGs), typically integrates empirical knowledge as free from bias as possible with the preferences of patients [20]. However, implementing and adopting recommendations within CPGs can be protracted [21] due to clinician perceptions [22], the volume and quality of the evidence [23], and even difficulties to integrate the mandates of different CPGs [17]. Specifically, previous findings from our group suggest that the decision-making of nurses was suboptimal regarding the adoption of CPG recommendations for preventing infectious complications and failure related to PIVC, highlighting behavioural and organisational differences between hospital environments and services [19]. Further exploring individual motivations, barriers and facilitators within organisations would contribute towards understanding the contextual elements that underpin decision-making around PIVC care [24,25]. Therefore, the purpose of this study was to investigate the determinants of suboptimal decision-making among nurses in Spanish hospital wards for the prevention of PIVC-related adverse events including PIVC failure and PIVC-BSI.

## METHODS

### Study Design and Setting

We conducted a qualitative study using semi-structured interviews to elicit perceptions, attitudes and beliefs about individual, team, and structural determinants of suboptimal PIVC management and care. The findings would strengthen the development of interventions and strategies to foster the implementation and diffusion of evidence-based recommendations in the health care service of Balearic Islands (Spain) [26].

We conducted the study in three hospitals. Hospital Manacor and Hospital Comarcal de Inca are state-funded acute care hospitals and serve a population of 150.000 and 130.000 inhabitants. These hospitals have 224 and 165 beds respectively, treating patients from all clinical specialities except cardiac, thoracic, and neurology surgery. The 3rd hospital, Hospital San Joan de Deu, is a state-funded, long-term care hospital with 197 beds mainly allocated to patients with chronic health problems or palliative needs.

### Participant Selection and Recruitment

Hospital ward nurses working routinely with inpatients at any of the three hospitals were considered for enrolment in the study. We approached key informants to identify suitable participants, who then in turn recruited other participants through a ‘snowball’ technique. We were keen to include participants from a variety of professional backgrounds and career pathways with a view to exploring rich experiences of managing PIVCs.

Participation was voluntary and without monetary compensation. A total of 28 individuals were approached, 19 (68%) of whom agreed in principle to participate, and with 14 participants finally interviewed. Selection, recruitment, and interviews ceased once data saturation was achieved.

### Data Collection

All semi-structured, face to face interviews were conducted by three researchers (two nurses and one psychologist). The interview guide used by Castro-Sánchez et al. [16] was adapted to the Spanish context, supplemented by a systematic review of the literature. The semi-structured interview (**Supplementary material 1**) was piloted in May 2018 to aid interview procedure and ensure an unambiguous understanding of the questions. The interviews were scheduled to last ∼45 minutes and conducted between September–November 2018 in locations and times convenient for participants. Field notes were made during and after the interview.

## Data Availability

The data that support the findings of this study are available on request from the corresponding author. The data are not publicly available due to privacy or ethical restrictions.

## Supplementary material 1. Interview guide

The interviews were audio-recorded and transcribed *verbatim*, with answers anonymized before the analysis. The transcripts were also returned to participants for comments and clarifications. An initial coding framework was applied to the interviews, which were then once more offered to respondents for validation of salient codes and themes.

### Data Analysis

The analysis of the data was carried out in a continuous and iterative manner aided by ATLAS.ti v7 software. In the inductive phase, transcripts were examined searching for units of meaning, and coded. These codes were grouped under broader categories and subcategories. Each transcript was independently coded by two researchers (HL-N and IB-M) who then met to compare their finding. During the deductive phase, data were analysed from the proposed elements of the theoretical framework and literature review.

We employed several triangulation strategies to underpin the methodological rigour of our analysis, following Guba and Lincoln’s approach [27]. Regarding methods, we compared the information collected in the recording of the interviews, the codes applied by both researchers, and the review by the participants to identify the coherence and any discrepancies of the discourse (member checking). In terms of data, two members of the research team shared and discussed their findings with each other. Another strategy to improve rigour was the meticulous development and recording of researchers’ reflectivity on any methodological decisions made throughout the study, as well as considering their dual status as clinicians and co-investigators [28]. Finally, the responses and initial analysis were discussed with two additional researchers with extensive experience in implementation science and qualitative research. The Consolidated Criteria for Reporting Qualitative Studies (COREQ) checklist was used. (**Supplementary material 2**).

## Supplementary material 2. The Consolidated Criteria for Reporting Qualitative Studies (COREQ) 32-item checklist for manuscript

### Research team and reflexivity

The knowledge about PIVC care and management by the research team was essential to interpret and contextualise the analysis. Three members of research team (two of which had previous experience in evidence implementation and vascular access research, and one in clinical psychology and social research), facilitated and conducted semi-structured interviews at the three participating hospitals. In addition, none of the researchers were linked with the participants, which allowed them to establish a rapport and fostered an open and frank discussion. The principal researcher is a doctoral candidate in a Translational Research in Public Health and High Prevalence Diseases programme.

### Ethical considerations

This study was approved by the appropriate research ethics committees. All participants were informed about the purpose of the study and their implications. We obtained write consent from participants.

## RESULTS

The interviews lasted an average of 35 minutes, providing rich data regarding the participants’ experiences about decisions on PIVC care to prevent the adverse events. Fourteen hospital ward nurses with a range of ages and clinical experience participated in the study. One nurse declined to participate in the study and two did not attend the scheduled interview. No reasons were given for non-participation. **Table 1** presents the characteristics of the participants.

**Table 1.** Demographics and professional characteristics of participants.

| Participant | Gender | Age (years) | Clinical experience (years) |
| --- | --- | --- | --- |
| 1 | Female | 41 – 45 | 16 – 20 |
| 2 | Female | 21 – 25 | 1 – 5 |
| 3 | Female | 31 – 35 | 11 – 15 |
| 4 | Female | 41 – 45 | 6 – 10 |
| 5 | Female | 41 – 45 | 16 – 20 |
| 6 | Male | 26 – 30 | 1 – 5 |
| 7 | Female | 26 – 30 | 1 – 5 |
| 8 | Female | 31 – 35 | 11 – 15 |
| 9 | Male | 41 – 45 | 6 – 10 |
| 10 | Female | 31 – 35 | 11 – 15 |
| 11 | Female | 21 – 25 | 1 – 5 |
| 12 | Male | 26 – 30 | 1 – 5 |
| 13 | Male | 26 – 30 | 1 – 5 |
| 14 | Female | 31 – 35 | 11 – 15 |

### Themes

Four major themes were identified related to determinants of care and are presented here together with illustrative quotations: 1) The ‘fog’ of decision-making in PIVC; 2) The ‘taskification’ of PIVC care; 3) PIVC care is accepted to be suboptimal, yet irrelevant; and 4) PIVC care gaps reflect behavioural shortcomings, yet proposed solutions only involve education and training.

The 15 codes emerging in our study (**Supplementary material 3**) indicated that these determinants were connected through PIVC care decision-making. **Figure 1** shows what determines optimal care of PIVCs among Spanish nurses.

**Figure 1.**
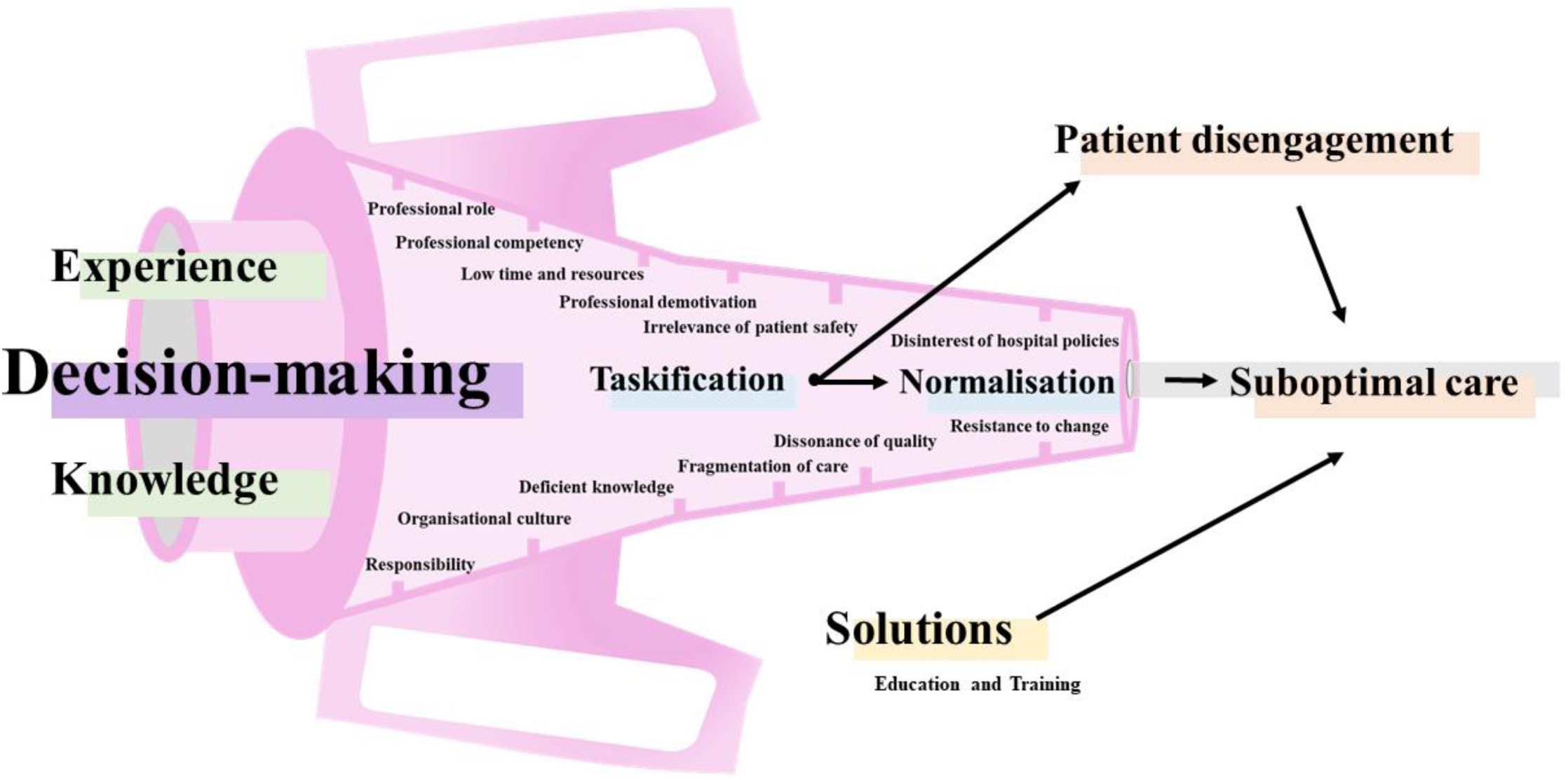
What fuels suboptimal care of peripheral intravenous catheters in Spanish hospitals?

## Supplementary material 3. List of Themes and codes

### 1) The ‘fog’ of decision-making in PIVC

Whilst nurses feel responsible for PIVC care, they do not however see themselves as the responsible decision-maker about PIVC insertion and removal. Such decisions are apportioned to physicians. But such demarcations of responsibility are not however clear cut, with frank ambiguities instead about some clinical decisions. Equally lacking is a jointly agreed upon framework of reference that explicitly allocates roles, professionals, and situations about PIVC care:

> ‘*I think it’s the nurse’s responsibility, the decision to insert a catheter, it’s the nurse’s as well. Well, or the doctor, because sometimes you cannot insert a PIVC, and the doctor decides to put in a central intravenous catheter*.’ Participant 1.

Such ambiguity is reinforced by servile relations between nurses and medical professionals, particularly in cases of urgency, and implicit delegation of tasks. This delegation appears to take place (or is not resented) only when nurses and physicians’ decisions are aligned:

> ‘*If it is a vital issue for the patient, we (nurses) can insert a vascular access without the authorization of the doctor. If you don’t [insert the vascular access], they [doctors] ask you why you haven’t inserted it. Yet sometimes when you do it, [insert the vascular access], they (doctors) tell you that you’ve exceeded your competencies*.*’* Participant 4.

### 2) The ‘taskification’ of PIVC care

Decisions about care, maintenance, management, and removal of PIVCs were highly fragmented and conducted as tasks rather than embedded within a nursing care process, resulting in a disjointed and often inefficient experience to reduce potentially infectious complications for patients. This *‘taskification’* may be defined as deficient knowledge of professional practices related to PIVC care, or dissonance between the perception of PIVC care offered and care effectively provided, where tasks rather than quality and safety healthcare are prioritised. One participant described this piecemeal approach to care:

> ‘*We (nurses) work routinely, performing tasks with automatic habits acquired from the ward. You observe the PIVC, disinfect it, you change the dressing and then you leave*’. Participant 1.
>
> ‘*We (nurses) have training. However, we lack an integral view about the management of care. Sometimes, even if you see that the PIVC is in perfect condition, if the patient says that it is hurting, then you should remove the catheter. Some kind of overall awareness is the key to better care*.’ Participant 5.

This focus on tasks rather than quality is exemplified by the perspectives of participants about the maintenance of PIVCs. Rather than another essential and valuable component of excellent PIVC care, nurses seemed only concerned about carrying it out appropriately to avoid wasting their time reinserting any catheters gone wrong, without a likely reflection on the patient experience or, even worse, implications towards healthcare-associated infections:

> ‘…*the interest in maintenance is quite low, what interests you as a nurse is that it takes you time not to do the technique (task) again, but not so much on the subject of infections*…’. Participant 9.
>
> ‘*Yes, there are failures, but not everyone fails in the same place. In maintenance people are less careful. People might fail to see a dirty PIVC or get wet because the patient has showered, and they don’t change it*…’ Participant 3.

Paradoxically, the interest in avoiding any waste of nursing time and resources was neutralised by decisions (or at least, aspirations) to ensure that all patients always had PIVCs, if possible. This blanket approach would appear to fit well with the taskification embedded in the continuum of care, as well as removing engagement for potential disputes with other professionals about the need (or lack of thereof) for PIVCs, an area fraught with uncertainty as previously highlighted:

> ‘*I don’t want a patient without PIVC, I don’t want to, I don’t feel safe. I have had some scares and if I can all patients would be carriers of a PIVC. If it were up to me, I would put everyone on a PIVC from the first day to the last day of hospital admission*.’ Participant 2.

### 3) PIVC care is accepted to be suboptimal, yet irrelevant

The reduction of PIVC care to an array of tasks surrounded by uncertainty resulting in suboptimal care for patients was acknowledged by the nurses, who tacitly accepted such status quo. Underpinning the inaction was the detachment from hospital policies and best practices, strengthened by their perceived flaws and ambiguity:

> ‘*The protocol is outdated and obsolete. I don’t think anyone has read it. For example, it recommends routinely changing PIVCs every 96 hours*…’ Participant 10.

These views about clinical practice guidelines as outdated and therefore irrelevant had further unwanted consequences. Often, the disinterest about the policies and the lack of motivation among nurses to adhere and uphold the mandates included in the protocols turned to active resistance against any changes in practice, and even anarchic behaviours:

> ‘*Yes, there is a hospital policy, but it’s kept in a drawer. No one looks at it, no one teaches about it, but we are expected to know that it is the standard of practice. But people do what they want*.*’* Participant 8.
>
> ‘*Perhaps if the hospital establishes a more accessible hospital policy, with clear and precise recommendations for the PIVC care*.’ Participant 11.

In addition, PIVCs were seen as having a low impact on patient safety during the management of intravenous therapy:

> ‘*In our ward we have a register of vascular access devices, where the day of insertion and maintenance is recorded. However, I don’t do it, I go to the patient and if I have to change the dressing, I do it and that’ s it*.’ Participant 4.

The haphazard approach to patient safety is reflected in some of the behaviours reported by the nurses, who for example recognise that covering the PIVC insertion site can lead to serious complications for patients, yet they frequently engage in that very same practice:

> ‘*About the issue of covering the catheter insertion site, many colleagues cover the site when they insert or maintain the catheter. This situation threatens patient safety, but we don’t care*.’ Participant 14.

A further dimension of this apparent insensibility to patient safety is the avoidance of patient preferences within decisions included in PIVC care:

> ‘…*It’s a relatively simple technique (patient education), which we have very internalized, it seems easy. However, it is difficult to comply with during PIVC care*’ Participant 9.

### 4) PIVC care gaps may reflect behavioural shortcomings, yet proposed solutions only involve education and training

Perhaps unsurprisingly, these failings are normalised as an inevitable consequence of structural deficits in education or training (‘*Nurses base their practice on what they learned in university or on day-to-day experience. They do not keep up to date or ignore the evidence*…’ Participant 10), or capital and human resources:

> ‘*Lack of time is a resource that hinders us to offer best care. This is nurses’ main complaint*’ Participant 6. ‘
>
> *Sometimes you find patients with a true PIVC ‘disaster’, perhaps due to lack of time or workload. Sometimes you don’t devote as much time as you would like to PIVC care*’. Participant 1.

The solutions offered by the participants were aligned with the gaps suggested, with *ad hoc* training (‘*Above all, we need training to nurses on the ward, even if they just were mini sessions. Ideally, they would be face-to-face or even online courses explaining how to manage and care for PIVCs*.*’* Participant 10) or specialised posts with leadership and expertise to mitigate and bring poor PIVC care to the fore:

> ‘*Role models are necessary to provide support, and make explicit the impact of professionals on PIVC, on the importance of optimal care management*.*’* Participant 4.

## DISCUSSION

Virtually every day nurses in all clinical settings engage with PIVC care, requiring knowledge and skills that include techniques such as catheter insertion, maintenance, and management, together with more social and behavioural skills such as negotiation and communication with other professionals, or patient education. Delving into the motivations and attitudes of nurses was essential to explore the determinants of decision-making about PIVC care (insertion, maintenance, management, and removal) to prevent complications such as PIVC-BSI and their consequences such as sepsis, ICU admission or even death for. Our study unearthed how the constellation of decisions associated with PIVC care was highly disjointed, resulting in an erratic pathway for patients and a suboptimal and wasteful process for the healthcare organiosation in the struggle against CRBSI.

In the Spanish context, the uncertainty and ambiguity reported in other settings [16] is further compounded by a taskification of PIVC care. Nurses outlined their daily PIVC workload along a dated, task-based, nursing model [29]. The cognitive impact of such PIVC task-stacking is not known [30], but the taskification may fuel and perpetuate a productivity fallacy whereby engaging in low- rather than high-value work is preferred, as the latter requires an individual and collective effort to assess not only what needs doing, but also what needs to stop [31]. Besides, nurses that fulfil delegated tasks with no explicit responsibilities and no clinical leadership could provide a sense of satisfaction and comfort during the shift. This stance would hinder nurses from efficiently managing care, as the short-term perspective about the tasks at hand would obstruct a longer-term, more efficient vision which would also be conscious of unnecessary resources used.

Healthcare professionals have traditionally perceived vascular access care as poorly related to patient safety, with more frequent adverse events are considered preventable [32,33]. This perception may justify the irrelevance of PIVC management and encourage the omission of care reported by our participants, individually and collectively, as a rational approach; driven by scarce resources, nurses would opt out of caring for the valueless PIVCs.

Our study does not offer any insight into the views of nurses on patient involvement in shared decision-making related to PIVC. However, participants acknowledged that they did not engage in patient education, arguably the initial requirement for patient implication in care. The consequences of such lack of engagement are not surprising, as seen in other studies reported by our group where we identified that ∼50% of patients did not know anything about the catheter they carried [19]. These findings are concerning in themselves, but also highlight the missed opportunities to embed patient education about multiple related safety areas such as infection prevention and control, hand hygiene, and vascular catheter care, where patients could have a crucial role [34].

Paradoxically, the interventions advocated by the participants to improve their practice focus on mitigating material deficits, but it is unclear how the increased resources would shift the nurses’ view on the impact of PIVCs on patient safety. The proposal for specialist nurses or vascular access specialist teams could improve the quality of the initial insertion and perhaps management of PIVCs [35], but risks, on the other hand, marginalising the interest of general nurses towards PIVCs even more. Furthermore, the lack of context-related knowledge may be other of the transcendent challenges to guarantee optimal practice in the healthcare system [36]. This scenario leads us that question the efforts made to improve the quality of interventions and the fidelity of the evidence-based practice [37].

Our findings contributed towards understanding the contextual information of different organisational environments as a baseline within our quality and safety improvement strategy related to PIVC care [38]. In this sense, exploring the nursing perceptions related to PIVC care could provide insights into how healthcare professionals construct their decision-making and the core components and contribute to the successful implementation process into clinical practice.

Our study presents some limitations. Our findings are clearly bound to the sociocultural, clinical, and organisational characteristics of the Spanish health and social care system, and the roles explicitly allocated to and implicitly claimed by nursing professionals. We carried out interviews with frontline nurses, and their perceptions of higher-level determinants (i.e., organisational arrangements) may not truly reflect existing institutional policies. Nonetheless, the strength of our study includes that exploring the determinants of optimal PIVC decision-making and management as experienced and constructed by nurses would enable the development of tailored quality improvement interventions [39]. Understanding contextual features is a first required step before effective knowledge transference at different levels [40,41]. To further understand the interaction between PIVCs policies and stakeholders, we plan to carry out a follow-up study with managers and decisionmakers which will provide contextual insights of meso and macro levels and elicit crucial information on barriers and facilitators.

## CONCLUSION

In conclusion, our study suggests exploring the determinants of suboptimal decision-making on preventing PIVC-BSI is vital. Uncertainty of responsibility, fragmentation of care coupled with a perception of low risk on the impact and quality of PIVC care can fuel a lack of adherence to recommendations for reducing infectious complications, disempowering patients in their self-care and ultimately harming patient safety.

The implementation of facilitation strategies, including the decision-making about determinants of PIVC care, could improve the adherence to best available evidence, raising awareness among nurses of the impact that care excellence has on patients’ health outcomes.

## List of abbreviations

PIVC: Peripheral intravenous catheter;
CRBSI: Catheter-related bloodstream infection;
PIVC- BSI: Peripheral intravenous catheter-related bloodstream infection;
CPG: Clinical practice guideline

## Ethical approval

The research ethics committee of Hospital Manacor and Balearic Islands approved this study (IB3794/18PI). All participants were informed about the purpose of the study and their implications. We obtained written consent from participants.

## Consent for publication

This manuscript does not contain data from any individual person.

## Authors’ contributions

IB-M is the principal investigator of the study. All authors contributed to the original idea, design of the study, and are responsible for the conduct of the study. IB-M and EC-S prepared the first draft of the manuscript. IB-M, JDP-G and EC-S conducted the qualitative analysis. All authors have confirmed their authorship in the document of responsibilities of the author. All authors provided critical commentary on drafts and approved the final protocol manuscript.

## Acknowledgements

The authors wish to thank all nurses for their participation. We would also like to thank the hospital managers and directors of nursing t Hospital Manacor, Hospital Comarcal de Inca and Hospital Sant Joan de Deu for their support.

## Twitter

Ian BLANCO-MAVILLARD, @IanBlanco7

Enrique CASTRO-SÁNCHEZ, @castrocloud

Gaizka PARRA-GARCÍA, @gaizka_pg

Miguel Ángel RODRÍGUEZ-CALERO, @MA_Rguez_Calero

Miquel BENNASAR-VENY, @miquelbennasar

Ismael FERNÁNDEZ-FERNÁNDEZ, @isma_Fdez

Joan de PEDRO-GÓMEZ, @JoanDePedro

