## Supplementary tables for "WHAT FUELS SUBOPTIMAL CARE OF PERIPHERAL INTRAVENOUS CATHETER-RELATED INFECTIONS IN HOSPITALS? – A QUALITATIVE STUDY OF DECISION-MAKING AMONG SPANISH NURSES": Appendix A1. Interview Guide.docx

SUPPORTING TABLE 1. INTERVIEW GUIDE

**Background of the nurse**

1. Gender

2. Age (years)

3. Education

4. Clinical experience

5. Type ward and clinical experience into the hospital ward

**Experience**

1. Can you tell me about your expertise regarding the peripheral intravenous catheter (PIVC) care?

**Principles of PIVC care**

1. Who do you think is responsible for the PIVC care (insertion, maintenance, management and education) of the PIVC?

2. How important do you perceive these interventions for the care of the PIVC within the care practice?

3. How do you perceive the evaluations and audits regarding your decision-making in the PIVC care?

4. How do you think health professionals can improve adherence to PIVC care recommendations?

**Barriers**

1. What holds you back from carrying out what you consider the best practice for the PIVC care?

2. What makes you feel that you couldn’t carry out the best practice for the PIVC care?

3. What do you think was lacking in your daily environment?

**Facilitators or required support**

1. What kind of support do you wish you had when deciding on the care of PIVC?
