## Supplementary tables for "WHAT FUELS SUBOPTIMAL CARE OF PERIPHERAL INTRAVENOUS CATHETER-RELATED INFECTIONS IN HOSPITALS? – A QUALITATIVE STUDY OF DECISION-MAKING AMONG SPANISH NURSES": Appendix A2. COREQ Checklist.docx

**Supplementary File S2. The Consolidated Criteria for Reporting Qualitative Studies (COREQ) 32-item checklist for manuscript:** What fuels suboptimal care of peripheral intravenous catheter-related infections in hospitals? – a qualitative study of decision-making among Spanish nurses.

| **Domain 1: Research team and flexibility** | | | **Page** |
| --- | --- | --- | --- |
| Personal Characteristics | | | |
| 1 | Interviewer/facilitator | IB-M, GP-G and HL-N | 5 |
| 2 | Credentials | MSc and PhD student | 7 |
| 3 | Occupation | Nurses and Psychologist | 5 |
| 4 | Gender | Males |  |
| 5 | Experience and Training | Three members of research team, of which two (IB-M, GP-G) had previous experience in evidence implementation and vascular access research, and one in clinical psychology and social research (HL-N). | 7 |
| Relationship with participants | | | |
| 6 | Relationship established | No relationship was established prior to study commencement. | 7 |
| 7 | Participant knowledge of the interviewer | Participants were made aware of the researcher’s role through the Participant Information Sheet. | 7 |
| 8 | Interviewer characteristics | Discussed in limitations regarding gender and experience of interviewers. | 7 |
| **Domain 2: Study design** | | | |
| Theoretical framework | | | |
| 9 | Methodological orientation and Theory | Data were analysed using thematic analysis, which was informed by a critical realist. | 6 |
| Participant selection | | | |
| 10 | Sampling | We approached key informants to identify suitable participants, who then in turn recruited other participants | 5 |
| 11 | Method of approach | Snowball sampling technique. | 5 |
| 12 | Sample size | 14 participants | 8 |
| 13 | Non-participation | Five nurses did not participate in the study, who had previously confirmed their agreement to participate due to data saturation. Nine nurse declined to participate and two did not attend the scheduled interview. No reasons were given for non-participation. | 8 |
| Setting | | | |
| 14 | Setting of data collection | The interviews conducted in locations and times convenient for participants. | 6 |
| 15 | Presence of non-participants | Only two researchers and participant were present during the interview. | 5 |
| 16 | Description of sample | Characteristics of the sample are reported at the start of the results section and presented in Table 1. | 8 |
| Data collection | | | |
| 17 | Interview guide | Details of the interview schedule are provided in the Data Collection section. The full schedule is provided in the supplementary materials. | 6 |
| 18 | Repeat interviews | No | n/a |
| 19 | Audio/visual recording | Interviews were audio-recorded and transcribed verbatim by the interviewer. | 6 |
| 20 | Field notes | Field notes were made during and after the interview | 6 |
| 21 | Duration | Interviews lasted an average of 35 minutes (shortest = 27; longest = 42). | 7 |
| 22 | Data saturation | Data saturation is discussed in the Methods section. | 5 |
| 23 | Transcripts returned | The interviews were audio-recorded and transcribed verbatim, with answers anonymized before the analysis. The transcripts were also returned to participants for comments and clarifications. | 6 |
| **Domain 3: Analysis and findings** | | | |
| Data analysis | | | |
| 24 | Number of data coders | In the inductive phase, transcripts were examined searching for units of meaning, and coded. These codes were grouped under broader categories and subcategories. Each transcript was independently coded by two researchers (HL-N and IB-M) who then met to compare and contrast their finding. During the deductive phase, data were analysed from the proposed elements of the theoretical framework and literature review. | 6 |
| 25 | Description of coding tree | A coding table is provided in the results, supporting table 2 | 9 |
| 26 | Derivation of themes | The analysis of the data was carried out in a continuous and iterative manner | 6 |
| 27 | Software | Atlas.ti v7 software was used for analysis and data management. | 6 |
| 28 | Participant checking | Regarding methods, we compared the information collected in the recording of the interviews, the codes applied by both researchers, and the review by the participants to identify the coherence and any discrepancies of the discourse (member checking). | 6 |
| Reporting | | | |
| 29 | Quotations presented | Quotations are presented to illustrate each of the themes and are identified using participant age, and work unit. | 9-14 |
| 30 | Data and findings consistent | There was consistency between data presented and the findings/ conclusions, evidenced by quotations presented. | 9-17 |
| 31 | Clarity of major themes | Major themes are discussed in the Results section. | 9-14 |
| 32 | Clarity of minor themes | Two sub-themes are discussed in the Results section. | 9-14 |
