## Supplementary tables for "WHAT FUELS SUBOPTIMAL CARE OF PERIPHERAL INTRAVENOUS CATHETER-RELATED INFECTIONS IN HOSPITALS? – A QUALITATIVE STUDY OF DECISION-MAKING AMONG SPANISH NURSES": Appendix A3. List of Themes and codes.docx

SUPPORTING TABLE 2. LIST OF THEMES AND CODES

| **Themes** | **Codes** |
| --- | --- |
| The ‘fog’ of decision-making in PIVC | - Responsibility - Professional competency - Professional role - Organisational culture |
| The ‘taskification’ of PIVC care | - Fragmentation of care - Deficient knowledge - Dissonance between perception care offered vs effectively provided - Low time and resources of nurses - Irrelevance of quality and patient safety - Professional demotivation - Unimportance of patient engagement |
| PIVC care is accepted to be suboptimal, yet irrelevant | - Disinterest of hospital policies - Resistance to change - Low impact of PIVC on patient safety |
| PIVC care gaps reflect behavioural shortcomings, yet solutions proposed only involve education and training | - Education and Training |
